# Healthcare Unplugged: Disparities in Broadband Internet and Health Facility Access Among US Counties

**DOI:** 10.1101/2024.12.18.24319288

**Authors:** Khushi Kohli, Stephanie Wang, Cody Chou, Bhav Jain, Kavya M. Shah, Mahi Kohli, Edward Christopher Dee, Sandeep Palakodeti

## Abstract

Despite telehealth expansion, access may remain lacking in high-burden communities with limited brick-and-mortar health facilities. We aimed to i) examine the relationship between broadband internet access and health facility availability and ii) explore disparities in broadband access. Using data from the 2017 National Neighborhood Data Archive, we obtained data on population density of outpatient care centers, diagnostic labs, and nursing/residential care facilities for 3133 US counties. Broadband access and sociodemographic data were obtained from the 2020 Mapping Broadband Health in America Platform and 2022 American Community Survey. Two-sample t-tests and multivariable linear regressions quantified the association between broadband internet access and health facility density, adjusting for sociodemographic covariates.

Counties with low broadband access had 12% fewer outpatient care centers and 48% fewer diagnostic labs than counties with high broadband access (all P < 0.001). A 1% increase in county population without broadband corresponds with a decrease of 0.0451 outpatient care centers, 0.0237 diagnostic labs, and 0.0886 nursing/residential care facilities per 100,000 people (all P < 0.001). Counties with limited in-person health facilities face reduced access to broadband internet, particularly in rural, low-income communities. Expanding broadband infrastructure and health services in these regions is essential.

## INTRODUCTION

With the recent and rapid growth of telemedicine services in the United States, disparate access to broadband internet poses major health challenges, particularly in rural communities.(1) Rural America already faces formidable healthcare inequities — while 25% of Americans live in rural communities, only 10% of physicians practice there.(2) Consistent trends of frequent hospital closures, sparser healthcare facilities, and lack of insurance coverage further exacerbate healthcare inaccessibility in rural communities, associated with higher burdens of diseases such as obesity, diabetes, and cancer.(3,4) Telemedicine serves as a potentially promising solution to overcome the inadequacies of physical health facilities in rural areas by circumventing the need to commute long distances to access healthcare.

Improving healthcare outcomes in these regions relies heavily on telemedicine. By allowing patients to consult with healthcare providers remotely, telemedicine can reduce the need for travel thus saving time and money and making it easier for patients to receive timely care.(5) It can also provide access to specialists who might not be available locally. For chronic disease management, telemedicine can enable continuous monitoring and follow-up care. Regular check-ins via telehealth and remote monitoring help in identifying early symptoms and managing conditions like diabetes, hypertension, and cardiovascular diseases effectively, thereby reducing hospitalizations and emergency visits.(6)

Yet, the potential of telehealth to improve healthcare accessibility in rural communities is hindered by the requirement of broadband internet for many of its functionalities, including video visits, remote monitoring of medical devices, and nonsynchronous messaging.(7) Despite a 38-fold increase in telehealth usage post-pandemic, high-burden communities with reduced access to local outpatient health facilities may still face limited telehealth access. Access to broadband internet significantly influences the utilization of telemedicine services. Broadband internet has been expanding across the United States, yet significant gaps remain, particularly in rural and underserved areas. Current barriers include not just the lack of infrastructure but also issues like affordability and digital literacy, which prevent people from making full use of available services.(8)

In this study, we focus on three types of health facilities: outpatient care centers, diagnostic labs, and nursing/residential care facilities. These facilities were chosen because they play important roles in providing comprehensive healthcare services. Outpatient care centers offer primary and specialized care, diagnostic labs provide essential testing and screening services, and nursing/residential care facilities support long-term care needs. High broadband access allows these facilities to integrate telehealth strategies and solutions effectively. For example, outpatient care centers can conduct virtual consultations, reducing the need for in-person visits. Diagnostic labs can use telehealth to provide remote consultations and quick access to test results, improving the speed and accuracy of diagnoses. Nursing and residential care facilities can leverage telehealth for remote monitoring and consultations, ensuring continuous care without the need for frequent hospital visits.(9)

We hypothesize that regions with lower access to these facilities may also have lower access to broadband internet. If true, such communities would experience “dual disparities” in access to healthcare, facing barriers in accessing not only physical health services but also the benefits of telemedicine. This could lead to delayed diagnoses, poor management of chronic conditions, and ultimately worse health outcomes in these communities.

Thus, this study aims to i) correlate broadband internet access with the availability of various health facilities and ii) highlight rural-urban and income-based disparities in broadband access. By addressing these issues, we hope to underline the importance of targeted interventions that can bridge the gap in both internet connectivity and healthcare services, ultimately improving health outcomes for rural and low-income populations.

## METHODS

### Data Collection

#### Population Density of Health Facilities

Using the University of Michigan’s 2017 National Neighborhood Data Archive (NaNDA), we extracted data on the population density of outpatient care centers, diagnostic laboratories, and nursing/residential care facilities for 3,133 US counties.(10) In this database, outpatient care centers are defined to include substance abuse treatment centers, kidney dialysis centers, and community health clinics; diagnostic laboratories include facilities that provide services related to medical testing and imaging; nursing/residential care facilities include skilled nursing facilities, inpatient care hospices, assisted living facilities that provide full-time nursing care, and facilities offering residential intellectual and developmental disability care. However, facilities that primarily offer housekeeping and social services were excluded from the definition of nursing/residential care facilities. The densities of these facilities were reported per 1,000 residents in the NaNDA database and were subsequently converted to densities per 100,000 residents.

#### Broadband and Demographic Data

We obtained county-level data on the percentage of households without broadband access from the Federal Communications Commission’s (FCC) 2020 Mapping Broadband Health in America Platform.(11) This data was derived from the FCC’s February 2020 filings of Form 477 by internet service providers, which includes information on national broadband speed and coverage rates. We then stratified counties into two groups: a “high broadband access” group, in which the percentage of households without broadband was below the median threshold of 9.8%, and a “low broadband access” group, in which the percentage of households without broadband access was above the median threshold of 9.8%.

Finally, we obtained county-level demographic data, including urbanicity status, household poverty rate, employment rate, insurance coverage rate, and sex, age, and race distributions from the 2021 US Census Bureau’s American Community Survey.(12)

### Mapping

To visualize geographical trends in healthcare facility density, broadband access, and poverty rates across the country, we used the QGIS 3.36.1 geomapping software.(13) Specifically, we mapped i) the population density per 100,000 residents of outpatient care centers, diagnostic laboratories, and nursing/residential care facilities, ii) the percentage of households without broadband access, and iii) the percentage of households below the poverty line to the US counties outlined in the 2019 TIGER/Line Shapefiles.(14) We then created geographical heatmaps for each variable using the mapped dataset.

### Data Analysis

Using two-sample t-tests, we substantiated the association between broadband access and the population density of outpatient care centers, diagnostic laboratories, and nursing/residential care facilities, respectively. Moreover, we conducted multivariable linear regressions to quantify the association between i) broadband internet access and the density of each facility and ii) household poverty rate and the density of each facility, adjusting for all sociodemographic covariates described above. All data analysis was conducted in R Version 4.3.2.(15)

## RESULTS

Using heatmaps to visually represent the county-level geographical distributions of outpatient care centers, diagnostic labs, and nursing and residential care facilities across the United States, our analysis revealed a high density of all three types of facilities in the Northwest and Northeast regions of the US. We observed elevated densities of diagnostic labs and nursing facilities in the Southeast, suggesting regional variations in resource distribution. Notably, all three types of facilities exhibited a significant dearth in a vertical belt across the central United States that traverses the Midwest (**Figure 1A-C**). In addition to facility distributions, we visualized the percentage of households without broadband access. Consistent with the spatial patterns observed for healthcare facilities, we observed a distinct belt across the Midwest characterized by a higher percentage of households lacking broadband access (**Figure 1D**).

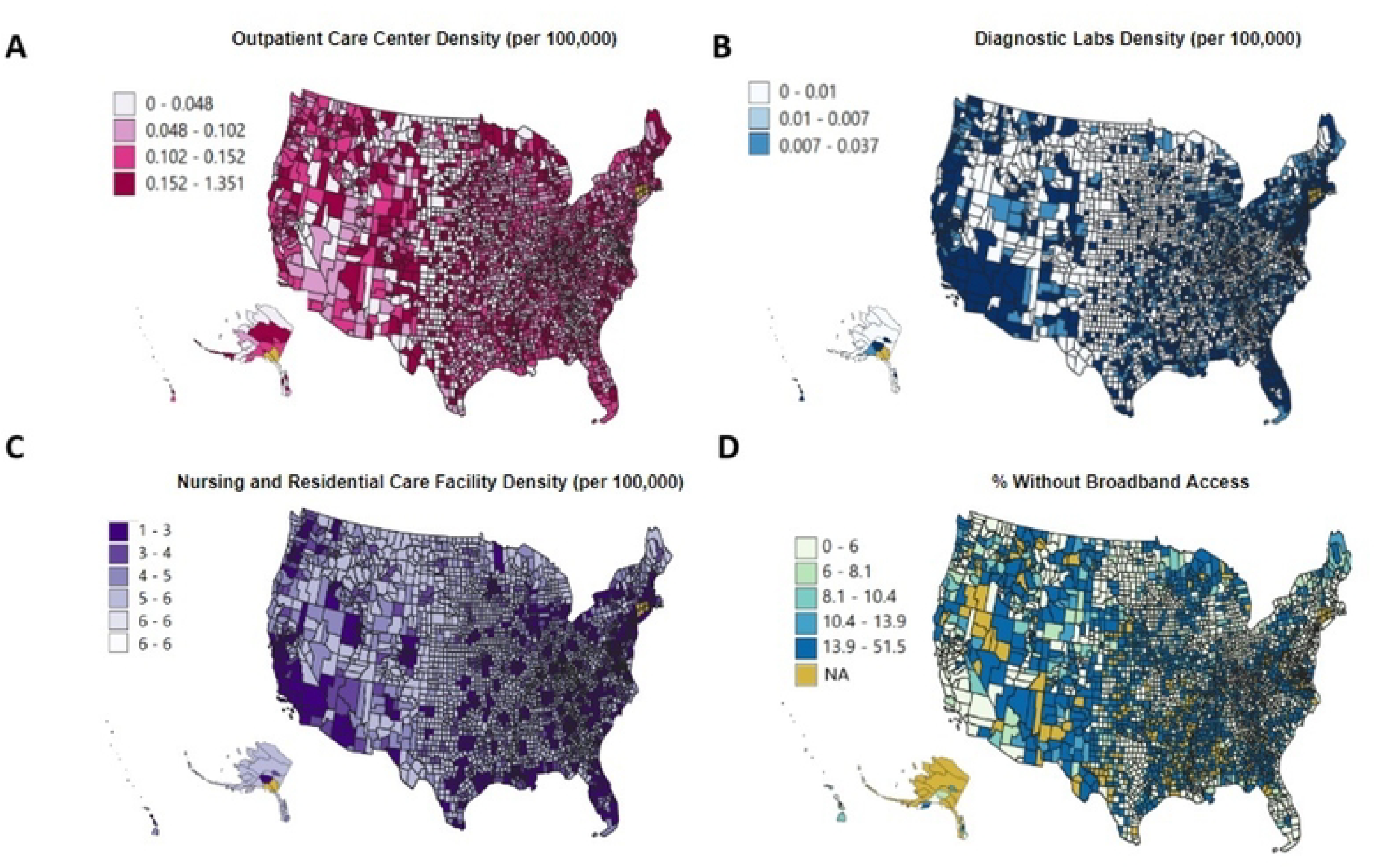

Two-sample t-tests revealed that US counties with reduced broadband access are significantly more likely to face reduced availability of various in-person health facilities. Low broadband access in US counties is linked to a 12% reduction in the density of outpatient care centers compared to counties with high broadband access (mean 10.46 vs. 11.91 outpatient care centers per 100,000 individuals, P < 0.001). Moreover, low broadband access in US counties was associated with a 48% reduction in the density of diagnostic labs (mean 1.91 vs. 3.95 diagnostic labs per 100,000 individuals, P < 0.001). However, broadband access did not show a statistically significant association with the density of nursing and residential care facilities (mean 31.36 vs. 30.66 facilities, P = 0.39) (**Table 1**).

**Table 1:** Population density of outpatient care centers, diagnostic labs, and nursing/residential facilities in US counties, stratified by access to broadband internet (two-sided *t*-test).

|  | Density (per 100,000 residents) |  |  | *P* value<br>(two-sided *t*<br>test) |
| --- | --- | --- | --- | --- |
| | All US<br>Counties | US counties with low<br>broadband access<br>(percentage of<br>households without<br>broadband access ≥<br>median) | US counties with high<br>broadband access<br>(percentage of<br>households without<br>broadband access $<$<br>median) | |
| **Outpatient Care Centers** | 11.18 | 10.46 | 11.91 | $P < 0.001$ |
| **Diagnostic Labs** | 2.93 | 1.91 | 3.95 | $P < 0.001$ |
| **Nursing/Residential Facilities** | 31.01 | 31.35 | 30.66 | $P = 0.39$ |

Furthermore, multivariable linear regression demonstrated that a 1% increase in the percentage of a county’s population without broadband access is linked to a decrease of 0.0451 outpatient care centers, 0.024 diagnostic labs, and 0.089 nursing and residential care facilities per 100,000 people (all P < 0.001). We also observed disparities in access to in-person health facilities along the dimension of income. A 1% increase in the percentage of households living below the poverty line is linked to a decrease of 0.25 outpatient care centers, 0.028 diagnostic labs, and 0.034 nursing and residential care facilities per 100,000 people (all P < 0.001) (**Table 2**).

**Table 2:**
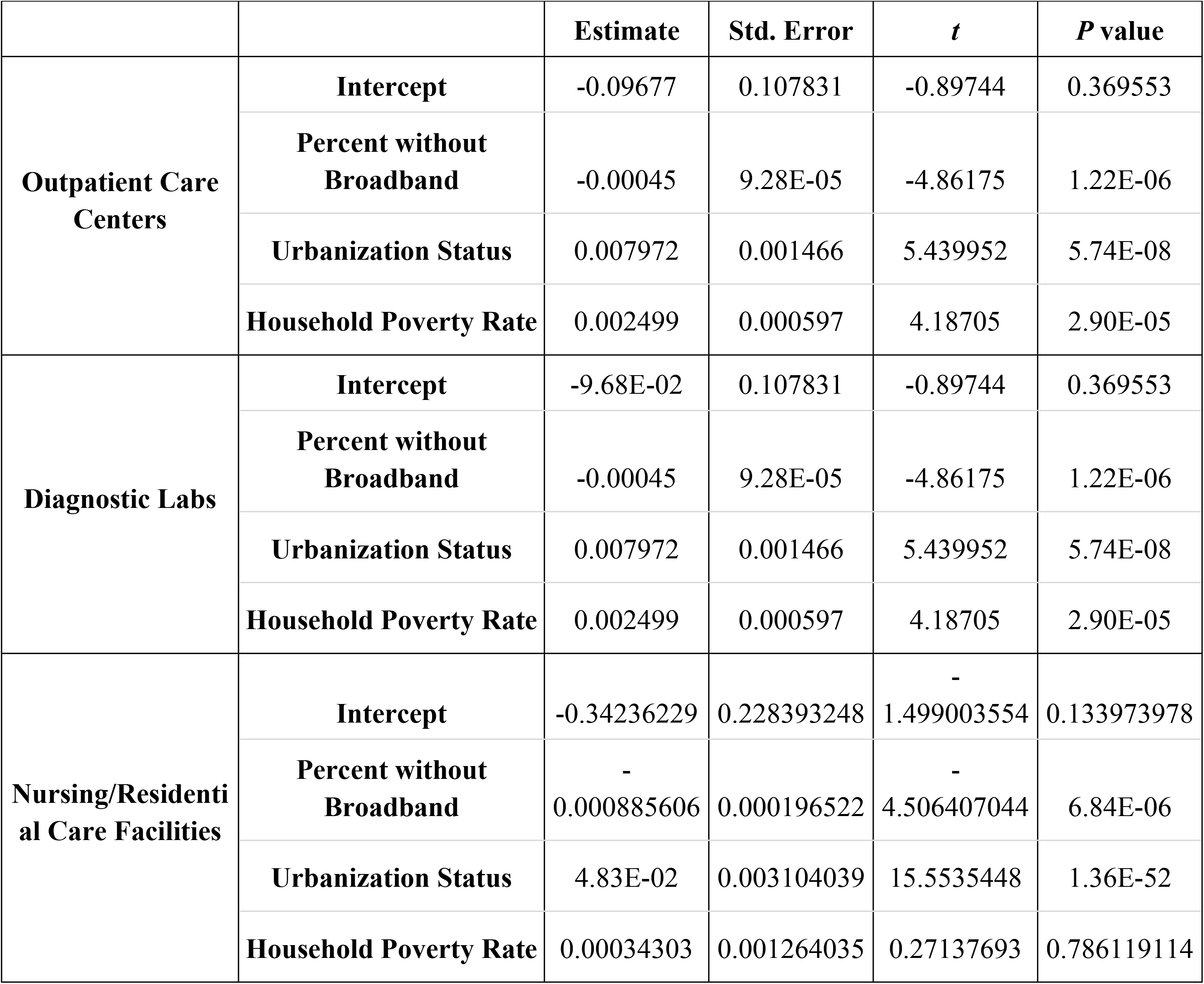
The effect of broadband access, urbanization status and household poverty rate on the population density of outpatient care centers, diagnostic labs, and nursing/residential facilities per 100,000 residents (two-sided *t*-test)

## DISCUSSION

Telemedicine has long been touted as a solution to rural-urban and socioeconomic disparities. However, in an era characterized by many post-pandemic telehealth interventions, rural communities with inadequate physical health facility presence may not benefit equitably from broadband internet-dependent telehealth services. Our results show a strong link between low broadband access and reduced availability of outpatient care centers and diagnostic labs, suggesting that rural and low-income communities may experience dual-disparities in access to both telehealth and in-person care.

Our findings align with previous studies that focus on the numerous sociodemographic disparities in broadband internet access. For example, previous studies have shown that rural populations generally face significant challenges in accessing specialty healthcare services. Due to the lack of specialists in rural areas, there is a greater reliance on primary care providers for conditions that often require specialized care which can delay early disease detection and complicate chronic disease management.(16) Often, people living in rural areas face long distances to healthcare facilities, higher out-of-pocket healthcare costs and limited insurance coverage, and the lack of critical healthcare infrastructure.(17) However, there has been limited focus on how broadband access may correspond with the distribution of these facilities. This study expands on the literature by examining the relationship between broadband internet access and the availability of various US outpatient care across the care continuum—including outpatient care centers, diagnostic labs, and nursing/residential care facilities. These facilities are critical because they provide essential services like early disease detection, diagnostic testing, and long-term care for chronic conditions—all of which are essential in ensuring continuous and comprehensive care. These findings shed light on the digital divide’s community-level impact on physical healthcare infrastructure, particularly in underserved rural and low-income communities, and highlights critical geographical and socioeconomic dual-disparities.

The connection between limited broadband access and reduced availability of healthcare facilities prompts the question of whether these services face lower demand in under-resourced areas. However, previous studies have shown that these areas actually have a higher demand for these services. In one study, people from rural communities often have a higher demand for healthcare services due to a greater burden of chronic diseases, such as diabetes, heart disease, and obesity. In addition, the study found that rural areas had significantly higher mortality rates from these chronic diseases compared to urban areas, including 193.5 vs. 161.7 deaths per 100,000 from heart disease, 180.4 vs. 158.3 from cancer, and 54.3 vs. 38.9 from chronic lower respiratory disease.(18) Our results are especially concerning because these populations face disproportionate health challenges, yet have reduced access to both broadband internet and in-person outpatient facilities.

To promote telehealth as a viable solution to combat rural health disparities, comprehensive community, clinical, and federal-level interventions, targeting both increased broadband access and physical health facility expansion, must be enacted. In the short term, initiatives to promote the development of broadband infrastructure may reduce disparities in healthcare access in rural communities. The FCC’s Affordable Connectivity Program (ACP) provided discounts on internet services and electronic devices to eligible households, with special provisions for households on Tribal lands. The ACP provided internet access to 21 million people, half of whom reported no or solely mobile internet service prior to their ACP benefit, with 80% reporting that affordability was the primary concern in having inconsistent or no service.(19) However, the ACP was discontinued in June 2024 due to a lack of congressional funding. One effective immediate intervention would be for Congress to reinstate funding for the ACP to support households that relied on it for broadband access, which were disproportionately low-income and Tribal households, including 15% from rural areas.(20) In tandem, given the underutilization of the ACP by eligible families during its implementation, it is important to raise awareness of programs like Link Health, which assists families in signing up for the ACP.(21)

Aside from publicly funded programs, eligibility and benefit expansion of private sector initiatives, such as T Mobile’s Project 10 Million can expand broadband access to underserved communities.(22) Project 10 Million aims to provide students qualifying for the National School Lunch Program with free internet and mobile hotspots and access to low-cost electronic devices. Expanding Project 10 Million provisions to geographic areas lacking in health facilities can assist families with accessing telehealth services, potentially bridging both health and educational disparities. Additional policies for improved telehealth access among rural and low-income communities include increased screening services, broadband infrastructure expansion, mandatory compensation for financial losses incurred during screening visits, and targeted investment in additional health facilities in high-burden regions, all of which are necessary to bridge the gap in access to early preventative care.

Furthermore, it is critical to address the shortage of in-person medical care in rural areas with limited access to telehealth services. Such efforts include increasing funding for rural health clinics to expand their staffing capacity, offering loan repayment and financial incentives to healthcare providers who commit to serving in rural areas, and implementing mobile health units to deliver essential medical services to remote regions. The MOBILE Health Care Act passed in 2023 expanded the development of mobile clinics through the use of federal funds, resulting in a 40 percent growth in mobile health center units since 2019.(23) Sustained and increased funding for this program can continue to incentivize the development of mobile health clinics for rural communities, increasing healthcare access. Additionally, policies should focus on expanding residency programs in rural hospitals, supporting the integration of telehealth into existing healthcare infrastructure, and creating partnerships between urban and rural healthcare systems to facilitate resource sharing and provider rotations.

This study bears several limitations. While providing valuable insights into broadband access and health facility density in US counties, the data does not include broadband service quality or post-2020 broadband accessibility data and thus may not fully capture post-COVID developments in infrastructure. Additionally, the reliance on county-level data may mask neighborhood-level variations and disparities, highlighting the necessity for a more granular investigation into factors influencing healthcare access in rural areas. Furthermore, our data does not consider broadband speeds, which remain an influential measure of the quality of broadband services beyond accessibility.

Finally, our study does not assess the relationship between broadband access and the density of emergency departments, which play a crucial role in providing immediate healthcare services, especially in rural or underserved areas.

Our research highlights a link between broadband inadequacies and scarcity of healthcare facilities in rural communities, underscoring the need for targeted broadband expansion in these areas as a critical strategy to enhance telehealth services and, by extension, improve healthcare access and outcomes. By investing in broadband infrastructure and attracting and retaining physicians in rural communities, policymakers can facilitate more effective delivery of telehealth services and address the root causes of healthcare disparities that disproportionately affect rural populations, ensuring that rural communities are not left behind in the digital age.

## Data Availability

All data was obtained from public repositories (described in Methods section).

